# Effects of High-dose Intravenous Vitamin C on Point-of-Care Blood Glucose Level in Septic Patients: A Retrospective, Single-Center, Observational Case Series

**DOI:** 10.1101/2020.11.23.20237461

**Authors:** Juan He, Guanhao Zheng, Xian Qian, Huiqiu Sheng, Bing Chen, Bing Zhao, Erzhen Chen, Enqiang Mao, Xiaolan Bian

## Abstract

**Introduction:** High-dose vitamin C (hdVC) is regarded as one of the essential adjunctive drugs for sepsis treatment. The present study aimed to deduce if hdVC could lead to erroneous testing results from the point-of-care glucose (POCG) measurement.

**Methods and materials:** This retrospective, single-center, observational case series involved septic patients treated by hdVC and monitored their paired POCG and laboratory glucose (LG) level for statistical analysis. The Parkes Consensus Error Grid Analysis was used as clinical influence assessment for paired blood glucose values. Subgroup analyses were conducted to explore the affecting extent of POCG readings by different VC dosage and various renal function level respectively.

**Results:** During the 3-year research period, 82 eligible septic patients who accepted at least three-day hdVC treatment were included in the current study. Compliance with ISO15197:2013 criteria was met in 30 (36.59%) paired values, which was far from the minimum criteria for accuracy. Subgroup analysis showed that worse renal function or higher VC dosage could lead to greater bias of POCG reading, but clinical risk would come forth scarcely while dealing with inaccurate POCG value.

**Conclusions:** High-dose intravenous ascorbate acid infusion could interfere POCG measurement values and LG method is more recommended, although significant medical risk hardly appears when physicians alter clinical action based on unreliable POCG reading.

## Introduction

Sepsis is a highly lethal clinical condition with systemic and inflammatory response to infection which could give rise to hemodynamic abnormality and multiple organ dysfunction syndrome (MODS)[1]. The basic treatment of sepsis includes anti-infectious treatment, source control, appropriate fluid resuscitation for maintenance of hemodynamic stability, as well as vasopressors[2]. Since ascorbic acid deficiency is widely observed and associated with rising morbidity in critically ill patients, high-dose vitamin C (VC) is regarded as one of the essential adjunctive therapies for treating sepsis because of its cost-efficiency and necessity[3]. Potential mechanisms of VC in sepsis treatment include amelioration of excessive oxidative stress and enhancement of cellular immune function [1, 4, 5]. The high-dose intravenous VC is considered to be a safe treatment, while only a few case reports have described the VC-associated calcium oxalate nephropathy as the primary adverse effect [2, 6].

As frequent blood glucose monitoring is essential to maintain normoglycemia for septic patients in intensive care unit (ICU) [7], controversy about hdVC interference of POCG measurement could not be neglected clinically. Erroneous blood glucose reading could cause clinical overtreatment about false hyperglycemia or hypoglycemia, which may bring on serious medical risk. Therefore, the current study focused on confirming whether hdVC could lead to erroneous testing results from POCG measurement as compared to the referential laboratory venous LG level measurement. The secondary aim of this study was to establish the correlation between different dosages of VC and the affected extent of POCG measurement level. Moreover, since elimination of VC in vivo mainly depends on glomerulus filtration and renal tubular reabsorption by sodium-dependent vitamin C transporter 1[8, 9], which may indicate VC is excreted by kidney principally, researchers of this study want to find out if renal function could be another key factor to affect the POCG result.

## Methods and materials

This retrospective, single-center, observational case series was conducted at the Emergency intensive care unit in Ruijin Hospital affiliated to Medical School of Shanghai Jiao Tong University, China. Because of the observational, non-interventional design, the inclusion of patients started after approval of the study by Ruijin Hospital Institutional Review Board and performed in accordance with the ethical standards of the Declaration of Helsinki 1964 and its later amendments or comparable ethical standards. VC was used for adjunctive therapy of sepsis in emergency intensive care unit (EICU) since 2013. The eligibility criteria of patients were as follows: (1) Age ≥18 years; (2) EICU admission between January 2016 and December 2019; (3) Fulfilled the standard classification of The Third International Consensus Definitions for Sepsis with SOFA score ≥2 within the initial 24h of admission[10]; (4) Treated by hdVC with corresponding dosage regimen for at least 3 days without using insulin, acetaminophen, mannitol and dopamine during hdVC treatment.

Patients who met the standard criteria were placed on hdVC infusion with corresponding dosage which depends on their severity of sepsis. In the current study, daily dosage of VC was between 50mg/kg to 200mg/kg. The constant infusion rate at 1 g/h of VC was applied in all patients. Time of starting infusion is daily fixed at 9 am. Dosage was not adjusted during continuous renal replacement therapy (CRRT). The duration of VC therapy lasted for at least 3 days and was dependent on the progression of the patient’s illness.

To verify the accuracy of POCG measurement, patients in this study underwent paired POCG and LG simultaneous measurements in two points of time as 15 minutes before first dose hdVC infusion and a quarter after last dose hdVC infusion, respectively. Time between paired POCG and LG blood sample collection were controlled within 15 minutes. POCG testing was performed by Medisafe mini blood glucose meter (Terumo Corporation, Tokyo, Japan) using the glucose oxidase-peroxidase (GOD-POD) colorimetric method. The laboratory LG measurements were carried out by Beckman Coulter AU5800 Chemistry Analyzer (Beckman Coulter Inc., S. Kraemer Boulevard, CA, USA) using a hexokinase spectrophotometric method. The source of the blood sample was not recorded.

Baseline data and clinical information, including age, sex, height, weight, Acute Physiology and Chronic Health Evaluation (APACHE) II scores, Systemic Inflammatory Response Syndrome (SIRS) scores, history of diabetes mellitus, serum creatinine level, CRRT modes (if paired blood glucose collection was in CRRT therapy period), POCG level, laboratory LG level and clinical outcomes were recorded by the researchers when each patient was on admission within 24 hours. SIRS scores were defined as 4 criteria: (1) fever (>38.0□) or hypothermia (<36.0□); (2) tachycardia (heart rate >90 beats/min); (3) tachypnea (respiratory rate >20 breaths/min); (4) leukocytosis, leukopenia, or bandemia (white blood cells >1,200/mm^3^, <4,000/mm^3^ or bandemia ≥10%). Each criterion owned 1 score and Patients who got at least two scores were considered as occurrence of SIRS[11, 12]. The hemoglobin level and hematocrit (HCT) was also recorded when hdVC usage was begun. Continuous data was recorded as median (interquartile range, IQR) values.

The primary endpoint of current study was to evaluate the accuracy of paired POCG and LG reading as well as the clinical significance of differences among paired POCG and LG results during the hdVC infusion period. Difference of paired POCG and LG results were calculated based on the Relative Difference Percentage (%) = [POCG]-[LG]/[LG] ×100%) between POCG and LG results.

Wilcoxon’s matched pairs test was performed to find out if there were significant differences between POCG and LG readings before and after the hdVC treatment period by SPSS (IBM® SPSS® Statistics v26.0.0.0, Chicago, IL, USA). The comparison of relative difference percentage before and after the hdVC treatment period were analyzed with the Kruskal–Wallis test by SPSS as well. The data was expressed as mean ± standard deviation. P-values<0.05 was considered as statistically significant.

When it comes to the clinical impact of unreliable POCG result during hdVC infusion period, only paired blood glucose samples collected after hdVC treatment were included in analysis of this step. ISO15197:2013 criteria[13] was performed as judging standard. As for the compliance of the criteria, at least 95% of POCG result need to be in range of ± 15 mg/dl when referential blood glucose result is less than 100 mg/dl, or within ± 15% of the LG result if the blood glucose level is no less than 100 mg/dl[14]. Bland-Altman method was utilized to further investigate the measurement of agreement among all paired POCG and LG results by GraphPad Prism Version 8.3.0 (538)[15]. The Parkes Consensus Error Grid Analysis, which was operated by Blood Glucose Monitoring System Surveillance Program[16], was aimed to assess the clinical significance of difference[17]. This error grid was divided into 5 zones which symbolized corresponding degree of risk associated with inaccurate measurement results. Zone A meant difference had no effect on clinical action. Zone B represented altering clinical action that could change little or no effect on clinical outcome. Zone C signified altering clinical action that could be likely to affect clinical outcome. Zone D was known as altering clinical action that could cause significant medical risk and Zone E defined clinical actions that could have dangerous consequences.

Secondary endpoints are the accuracy of POCG measurement in patients based on different dosages and renal function. In our study, all patients were divided in two groups by dosage levels. Patients with higher dosage were administered VC from over 100mg/kg to 200mg/kg per day, while patients with lower dosage were daily treated by VC from 50mg/kg to no more than 100mg/kg. Categorization was carried out according to patients’ renal function as well. Paired blood glucose collection from patients with an increased creatinine clearance of greater than 130 mL/min/1.73 m^2^ on admission was classified as ‘Augment Renal Clearance (ARC)’. Paired blood glucose acquired and measured when patients underwent CRRT treatment was determined as ‘CRRT’. Patients who had a history of Chronic Kidney Diseases (CKD) or blood serum creatinine ≥ 150% patients’ baseline serum creatinine within 24-hour admission period defined as impaired renal function[18]. Correspondingly, non-impaired renal function was identified as patients’ blood serum creatinine ≤ 150% baseline serum creatinine within 24-hour admission period and lack of a history of CKD. Baseline serum creatinine was derived from patient’s personal electronic medical record. Creatinine level on admission was also considered as baseline serum creatinine only when patients’ baseline serum creatinine was lack of adequate information in their medical record.

## Results

From January 2016 to December 2019, 656 patients were consecutively admitted to the EICU in the hospital. Among 141 potentially eligible septic patients treated by hdVC during this period, 59 were excluded (10 patients’ age were under 18, 27 had insulin administration in the VC therapy period, 16 were treated with VC less than 72h, and 24 used dopamine as concomitant medication) and 82 (58.16%) fulfilled the criteria for inclusion in the present study (Table 1).

**Table 1.**
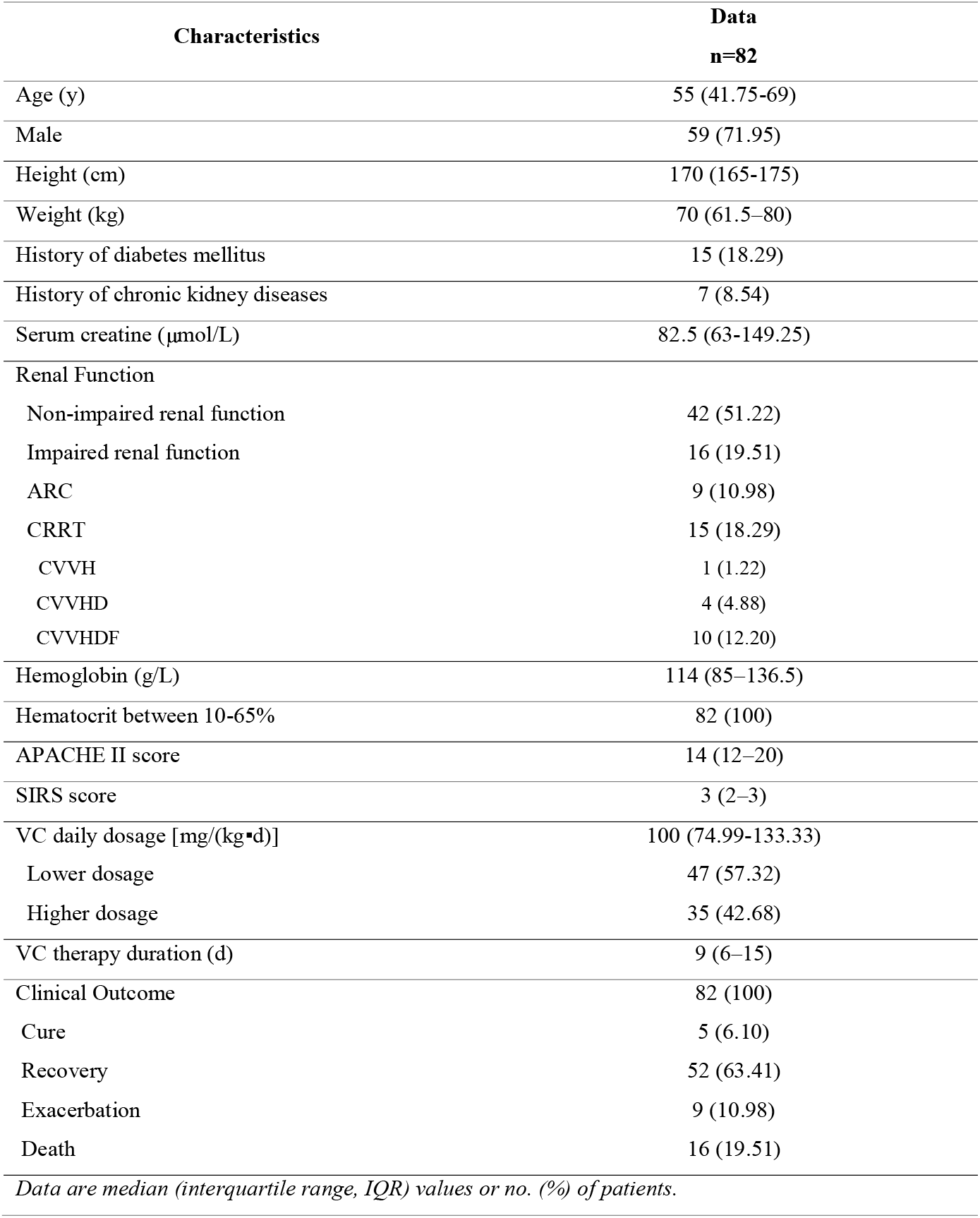
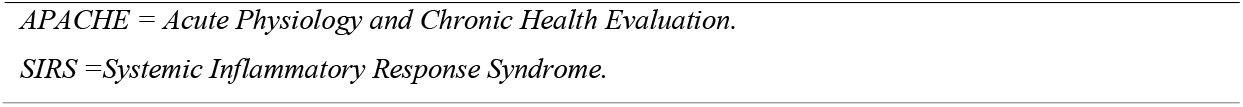
Characteristics of the 82 patients with VC therapy

Concomitant medication was a major influencing factor of POCG testing. Acetaminophen, dopamine, and mannitol might affect the POCG reading results, especially for dosage-dependent false-positive interference with glucose measurements by dopamine[19]. Insulin could cause blood glucose sharply dropping in a very short time. Therefore, patients who accepted acetaminophen, dopamine, mannitol and insulin treatment were excluded for reducing interference of the current study.

Furthermore, HCT interference was another crucial factor of POCG accuracy. The hematocrit range of 35 to 60% was considered as a reliable range for minimizing measuring bias [20, 21]. All 82 patients in the current study met this criterion and no one was excluded from the research.

The 82 patients displayed 14 (IQR 12-20) and 3 (IQR 2-3) for APACHE II score and SIRS score, respectively. Approximately, 18% of patients presented a medical history of diabetes mellitus. The median serum creatinine level of all patients was 82.5 (IQR 63-149.25). The hemoglobin level was recorded as median 114 g/L (IQR 85-136.5). Moreover, 15 patients (18.29%) had accepted CRRT during VC therapy period (mostly Continuous Veno-Venous Hemodiafiltration (CVVHDF) mode for 10 patients), and 16 patients (19.51%) had varying degrees of impaired kidney function. Normal kidney function and ARC accounted for 42 (51.22%) and 9 (10.98%) patients, respectively. Despite the positive and effective treatment in EICU, 16 patients succumbed to various kinds of refractory pathogenesis eventually and 9 patients were transferred out from EICU with exacerbating condition. Five patients cured totally from their own illness and 52 patients got recovered gradually. The median number of VC dosage was 100mg/(kg?d) (IQR 74.99-133.33) and VC therapy duration was 9 days (IQR 6-15). 35 (42.68%) patients were accepted higher dosage treatment and 47 (57.32%) patients were taken lower dosage therapy.

For all patients, POCG value was significantly lower than LG reading whether before hdVC infusion (158.08 ± 62.08 mg/dl vs 186.25 ± 71.50 mg/dl, *P*<0.001, Table 2) or after hdVC treatment (142.90 ± 58.91 mg/dl vs 186.74 ± 72.74 mg/dl, *P*<0.001, Table 2). Relative difference percentages from paired blood glucose of patients who had finished hdVC treatment were significantly higher than those of patients who had not started hdVC infusion(−17.86% ± 35.68% vs -12.43% ± 21.44%, *P*=0.01, Table 2), which implied that infusion of hdVC could increase the differences between POCG and LG values indeed.

**Table 2.**
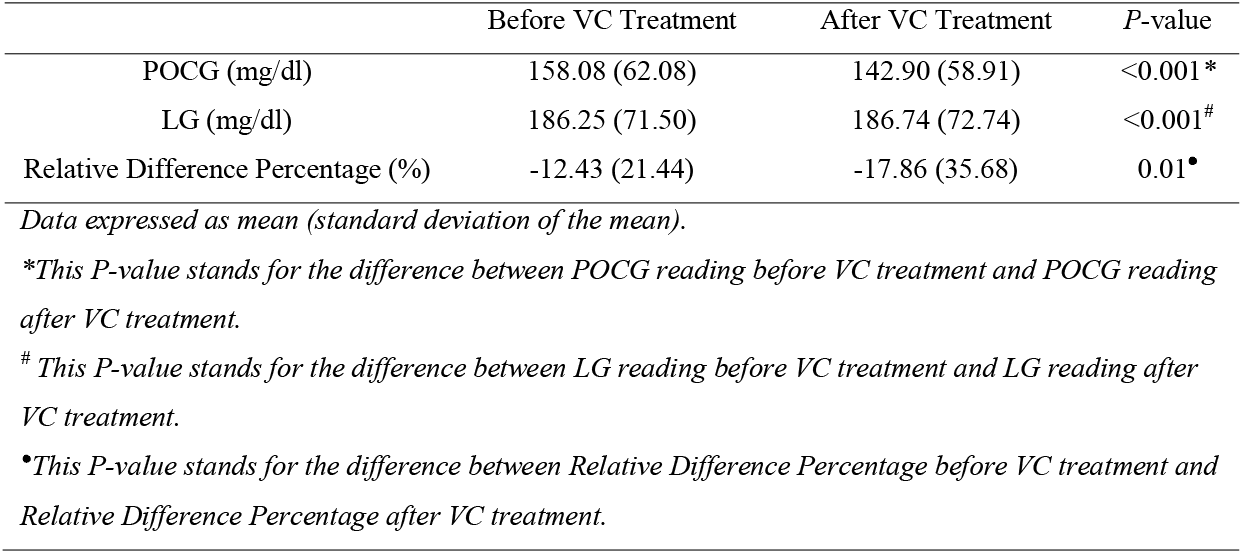
Statistical analysis among all POCG and LG paired values before and after VC treatment

The accuracy of POCG reading measurements compared to LG reading measurements was only 36.59% (Table 3), which indicated a failure to meet the minimum 95% accuracy standard following the compliance with ISO 15197:2013 criteria. The Bland-Altman plot (Figure 1) presented that mean difference between POCG and LG result was -43.83 mg/dl. The mean POCG values were obviously lower than mean LG readings with 95% limits of agreement of -177.97 and 90.32 mg/dl. The Parkes Consensus Error Grid Analysis (Figure 2) of POCG variation showed that difference of 2 (2.44%) paired blood glucose could induce a significant medical risk when altering clinical action. However, little clinical impact of POCG differences was exhibited under most circumstance since 42 (51.22%) paired blood glucose results fell into Zone A in the Parkes consensus error grid analysis, Zone B and Zone C owned 19 (23.17%) paired samples equally as well.

**Table 3.**
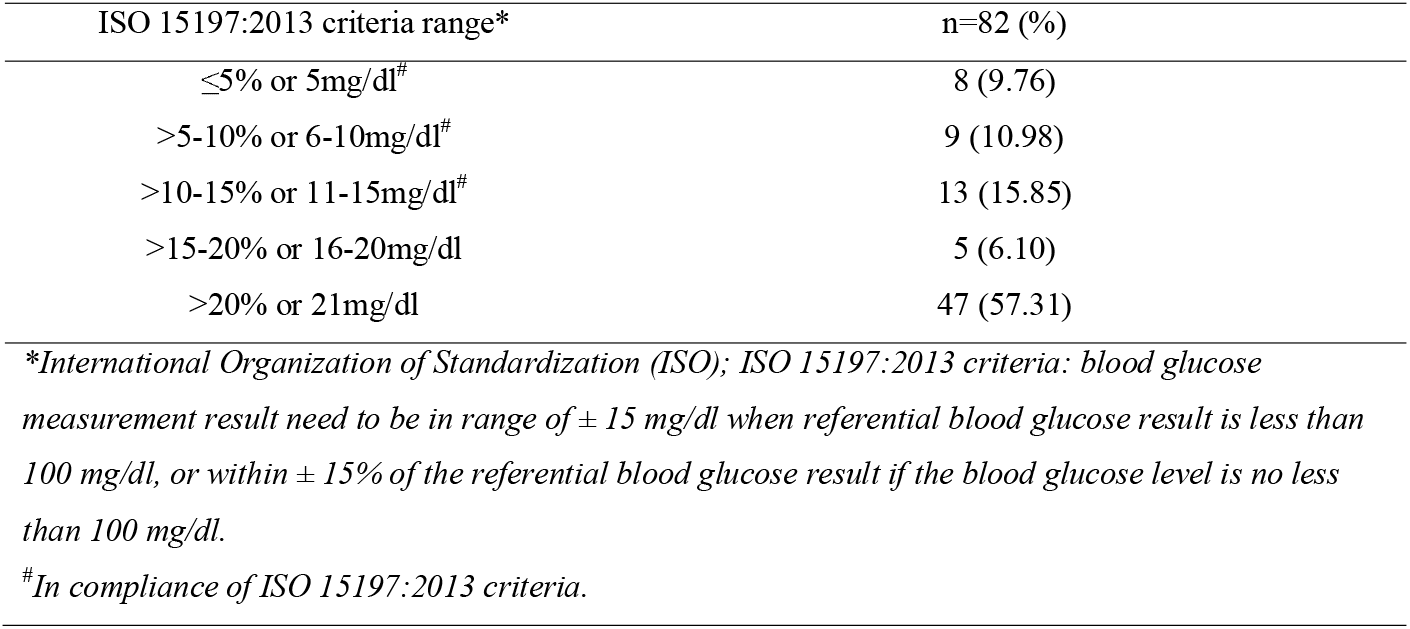
Difference of POCG compared to LG for all paired glucose values by ISO 15197:2013 criteria range

**Fig. 1.**
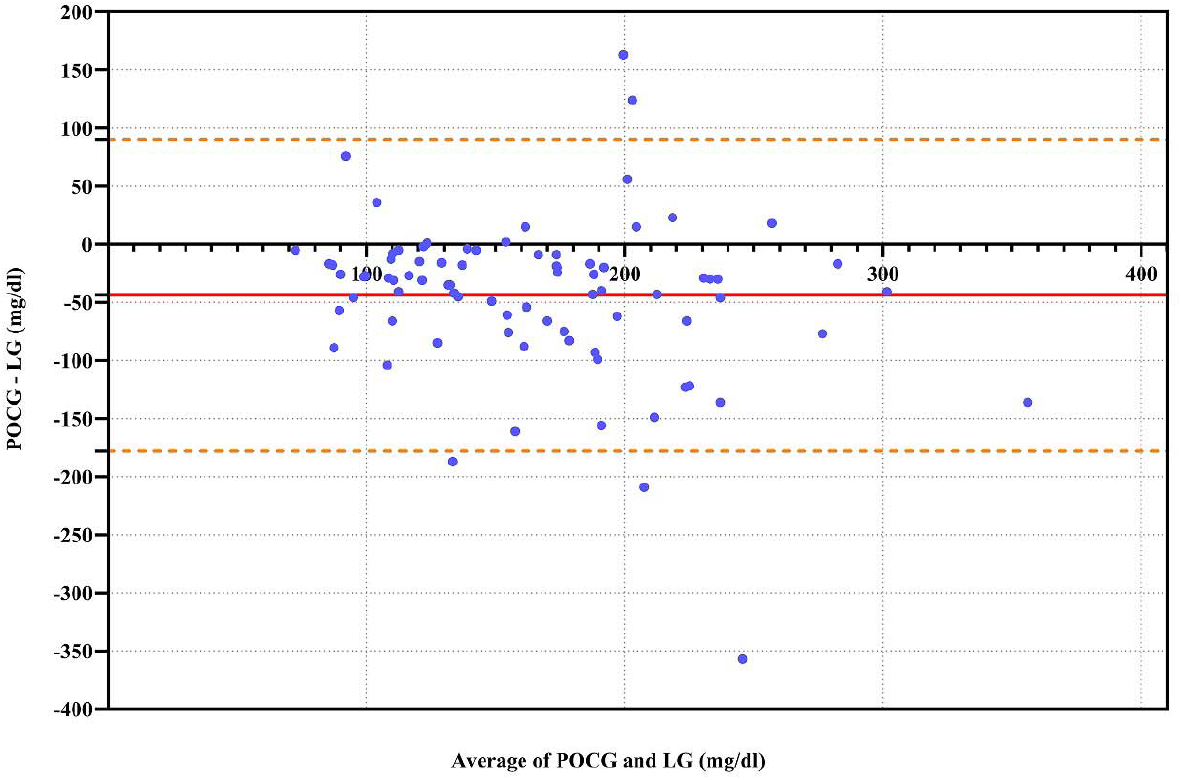
Comparison between POCG and reference LG levels by Bland-Altman plot. The red solid line means the mean difference among all paired data, namely bias. The orange dashed lines represent the 95% limits of agreement.

**Fig. 2.**
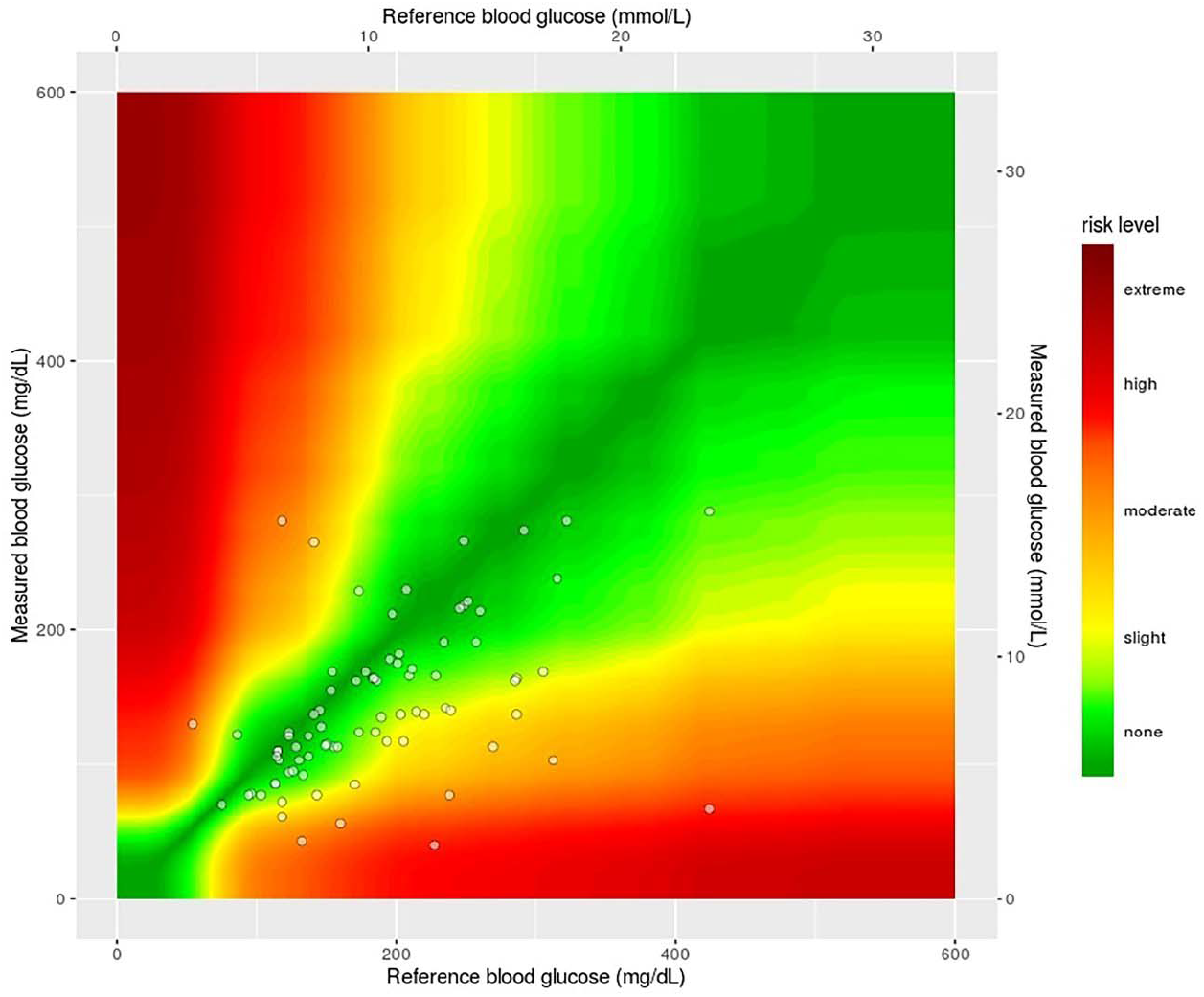
The Parkes Consensus Error Grid Analysis of paired blood glucose readings between POCG and LG. This error grid was divided into 5 zones which symbolized corresponding degree of risk associated with inaccurate measurement results. Zone A meant difference had no effect on clinical action. Zone B represented altering clinical action that could change little or no effect on clinical outcome. Zone C signified altering clinical action that could be likely to affect clinical outcome. Zone D was known as altering clinical action that could cause significant medical risk and Zone E defined clinical actions that could have dangerous consequences.

Dosage classification for paired blood glucose level with the compliance of ISO 15197:2013 criteria was detailed in Table 4. Forty-seven paired blood glucose values collected as lower dosage and only 18 (38.30%) pairs followed ISO 15197:2013 criteria. Compliance with ISO 15197:2013 criteria was met in 12 (34.29%) paired POCG and LG results of 35 evaluated as paired blood glucose collection of higher dosage. The Parkes Consensus Error Grid Analysis showed that 76.6% and 71.42% blood glucose pairs from lower and higher dosage group fell into Zone A and Zone B respectively, while 5.71% blood glucose pairs from higher dosage caused inaccurate POCG measurement results that could significantly affect clinical outcome. No Paired blood glucose result from lower dosage group included in Zone D and Zone E (Table 5).

**Table 4.**
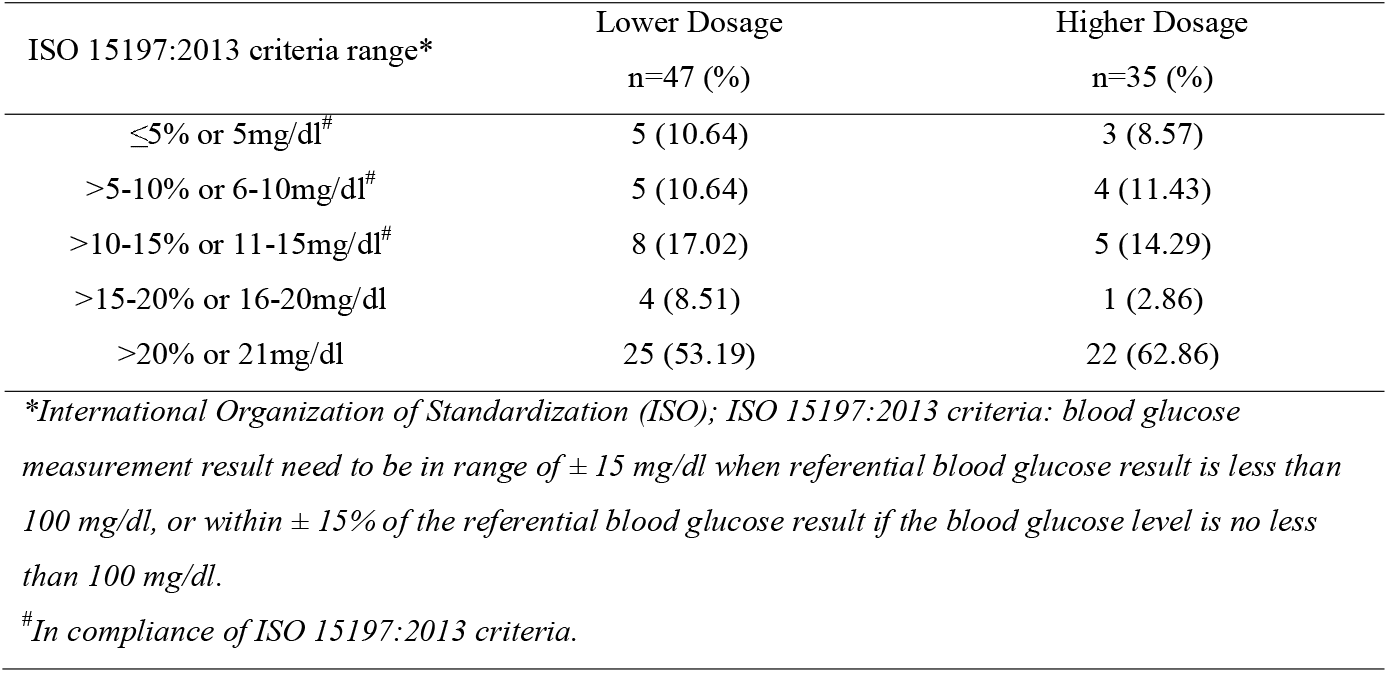
Subgroup Analysis for various dosage about difference of POCG compared to LG for all paired glucose values by ISO 15197:2013 criteria range

**Table 5.**
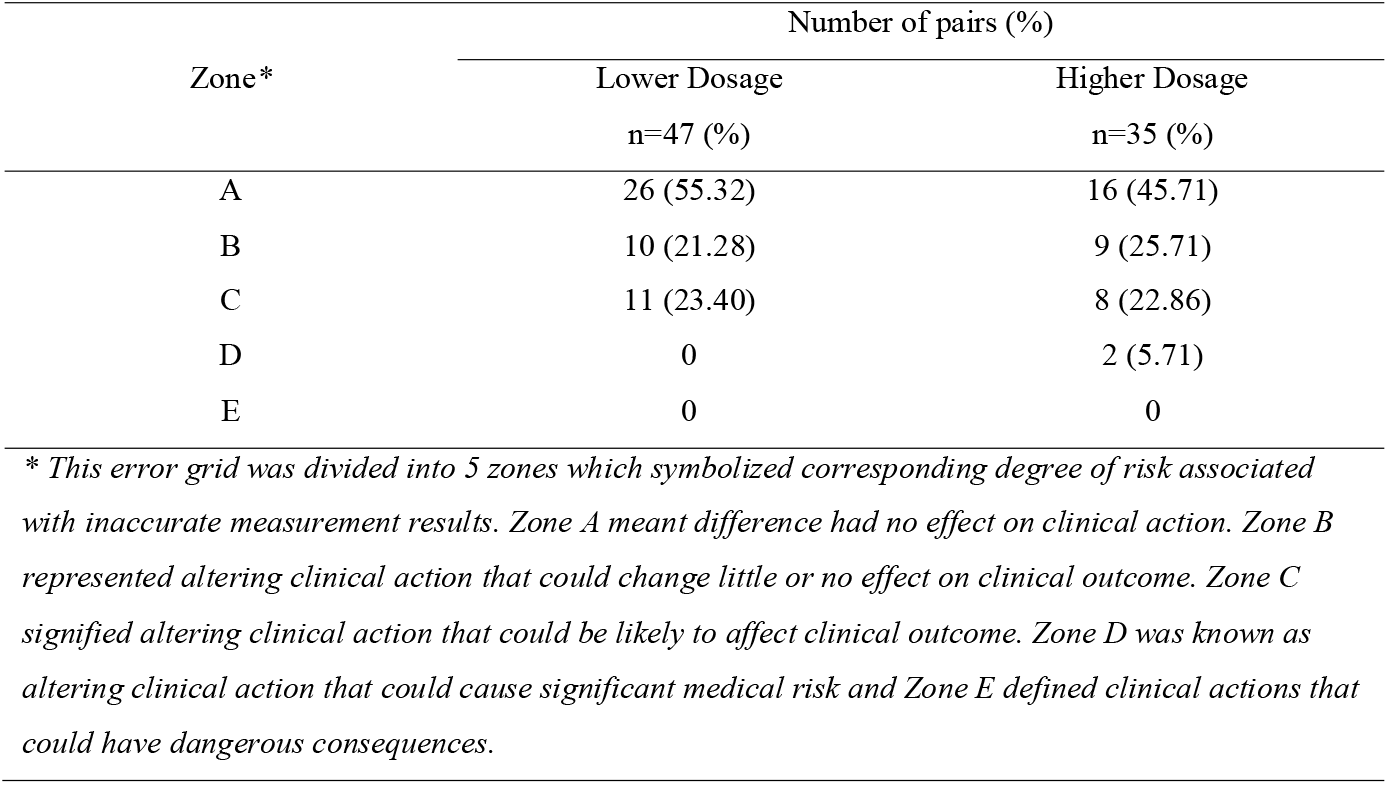
The Parkes Consensus Error Grid Analysis for POCG values compared to LG readings for various dosage groups

Furthermore, renal function was another key stratified factor for all paired blood glucose collection in this research. There were 42 patients with non-impaired renal function. Nineteen (45.24%) of them provided paired blood glucose readings in compliance with ISO 15197:2013 criteria. Six (37.5%) paired blood glucose values among 16 paired blood glucose measurement from patients with impaired renal function met the criteria. With regards to the rate of compliance about aforementioned criteria, only 22.22% and 20% for CRRT and ARC patients respectively (Table 6). Table 7 exhibited the result of Parkes Consensus Error Grid Analysis for each renal function level. Blood sample pairs from patients with non-impaired renal function (78.58%) or augment renal clearance (88.89%) had higher possibility to categorize in Zone A and Zone B than those from patients with impaired renal function (68.75%) or suffering CRRT treatment (60%). In addition, both impaired renal function and CRRT group owned one paired blood sample which fell into Zone D.

**Table 6.**
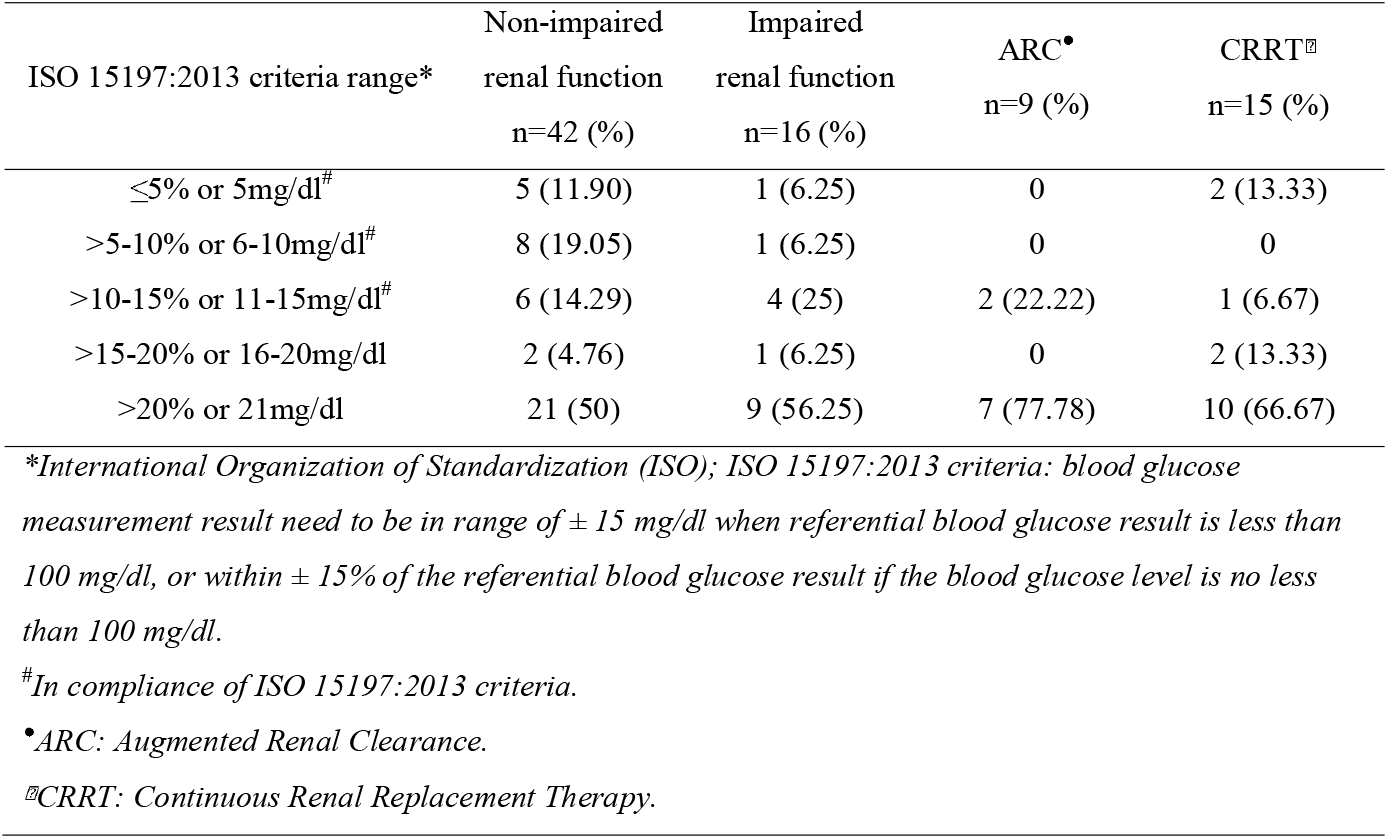
Subgroup Analysis for diverse renal function about difference of POCG compared to LG for all paired glucose values by ISO 15197:2013 criteria range

**Table 7.**
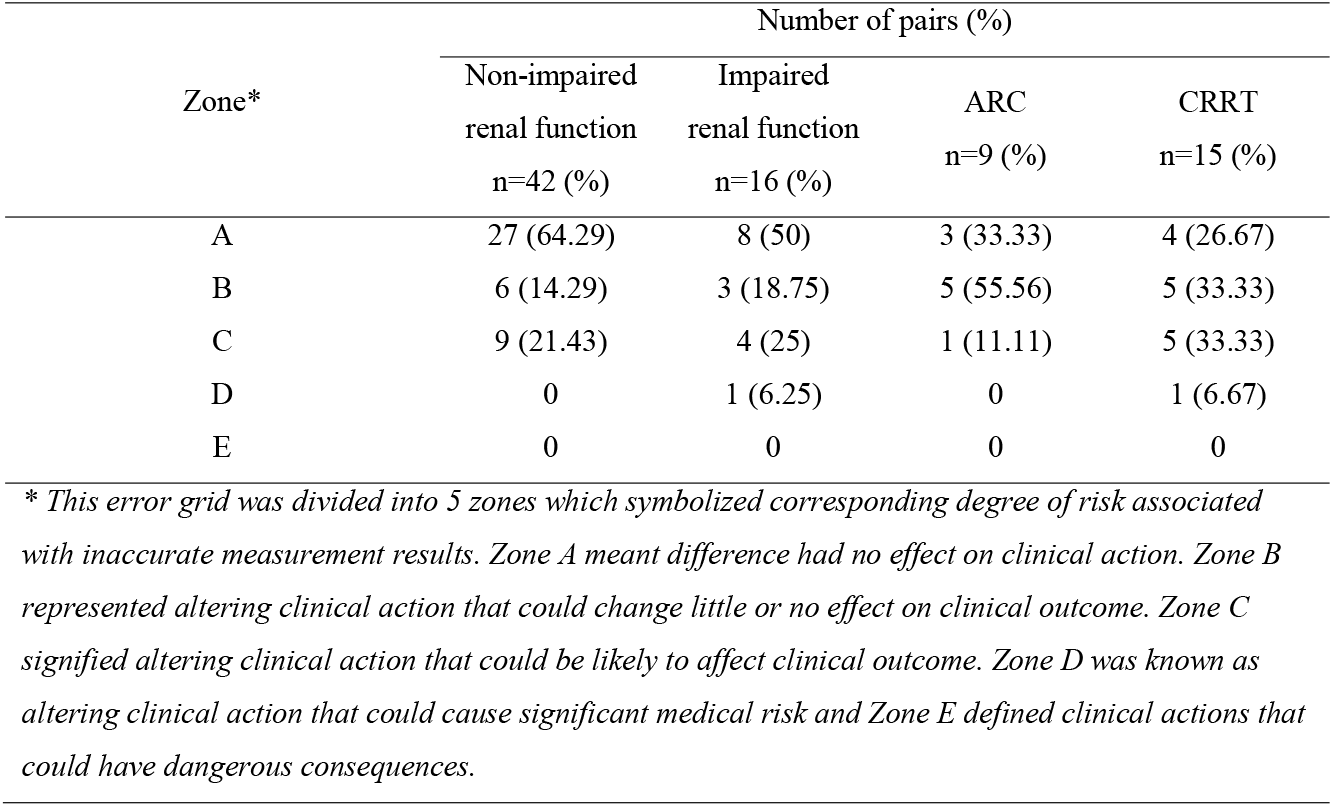
The Parkes Consensus Error Grid Analysis for POCG values compared to LG readings for groups of different renal function level

## Discussion

Clinically, drug interference of POCG value would lead to the inappropriate therapy for fictitious hyperglycemia or hypoglycemia[22]. Improper treatment with hypoglycemic agents for fictitious hyperglycemia could bring about a series of clinical symptoms like trembling, heart racing, nausea and sweating, even coma or death in some severe cases[23]. False low POCG reading might induce practitioners using high concentration of glucose supplement for patients, which could elevate blood glucose level above normal value. High blood glucose status usually accompanied with higher risk of infection, even increased mortality rates for septic patients[24].

High dose ascorbic acid causing inaccurate POCG reading is widely concerned clinically these years. According to the retrospective review by Kahn *et al*., 18 burn shock patients were administered hdVC (66 mg/kg/h × 18 h) infusions during burn resuscitation; 5 were chosen for the subsequent analysis. High-dose VC infusion for burn shock would give rise to fictitious hyperglycemia for POCG measurement as compared to the LG measurement result [25]. Sartor *et al*. designed another case series for the direct comparison of POCG and LG values in 7 patients with >30% burns on the total body surface area (TBSA); these patients received 66 mg/kg/h VC treatment. The study found that POCG level was elevated during the infusion period in all the patients [26]. Conversely, a prospective observational pilot study was designed by Smith *et al*., wherein 5 patients were administered VC at 1.5 g q6h/d, which did not lead to clinically significant differences between POCG and LG results [27].

Therefore, this study aimed to explore if high dose ascorbate acid infusion could cause fictitious value according to the POCG results as compared to the LG measurement. Moreover, the secondary target was to verify whether VC dosages or kidney function are the putative influencing factors with falsely readings of POCG.

In our research, statistical bias was detected about the measurement results from POCG using the GOD-POD method. There was a significant lower value for POCG compared to paired LG result after hdVC infusion. Statistically significant augmentation of relative difference percentages between POCG and LG reading was observed in period of hdVC post-infusion as well. This might imply that POCG measurement is unreliable when hdVC is used as adjunctive therapy, which suggested avoiding POCG as the blood glucose measurement when patients use hdVC treatment, besides LG is strongly recommended as the alternative ideal blood glucose monitoring method instead of POCG measurement.

A rational explanation of this phenomenon was that different methods were applied in POCG and LG testing, respectively. The GOD-POD colorimetric method is employed for POCG testing while the hexokinase method is for LG measurement.

The hexokinase spectrophotometric method is specific for measuring glucose in the serum or plasma (Figure 3a). Initially, hexokinase plus adenosine triphosphate (ATP) transforms glucose to glucose-6-phosphate (G-6-P) plus adenosine diphosphate (ADP), followed by a reaction started by G-6-P, NAD^+^, and G-6-P dehydrogenase to generate NADH, which is measured spectrophotometrically at 340 nm. NADH is proportional to the glucose level, and no interaction was observed between NADH and ascorbic acid. VC did not interfere with the results of the hexokinase method [25].

**Fig. 3.**
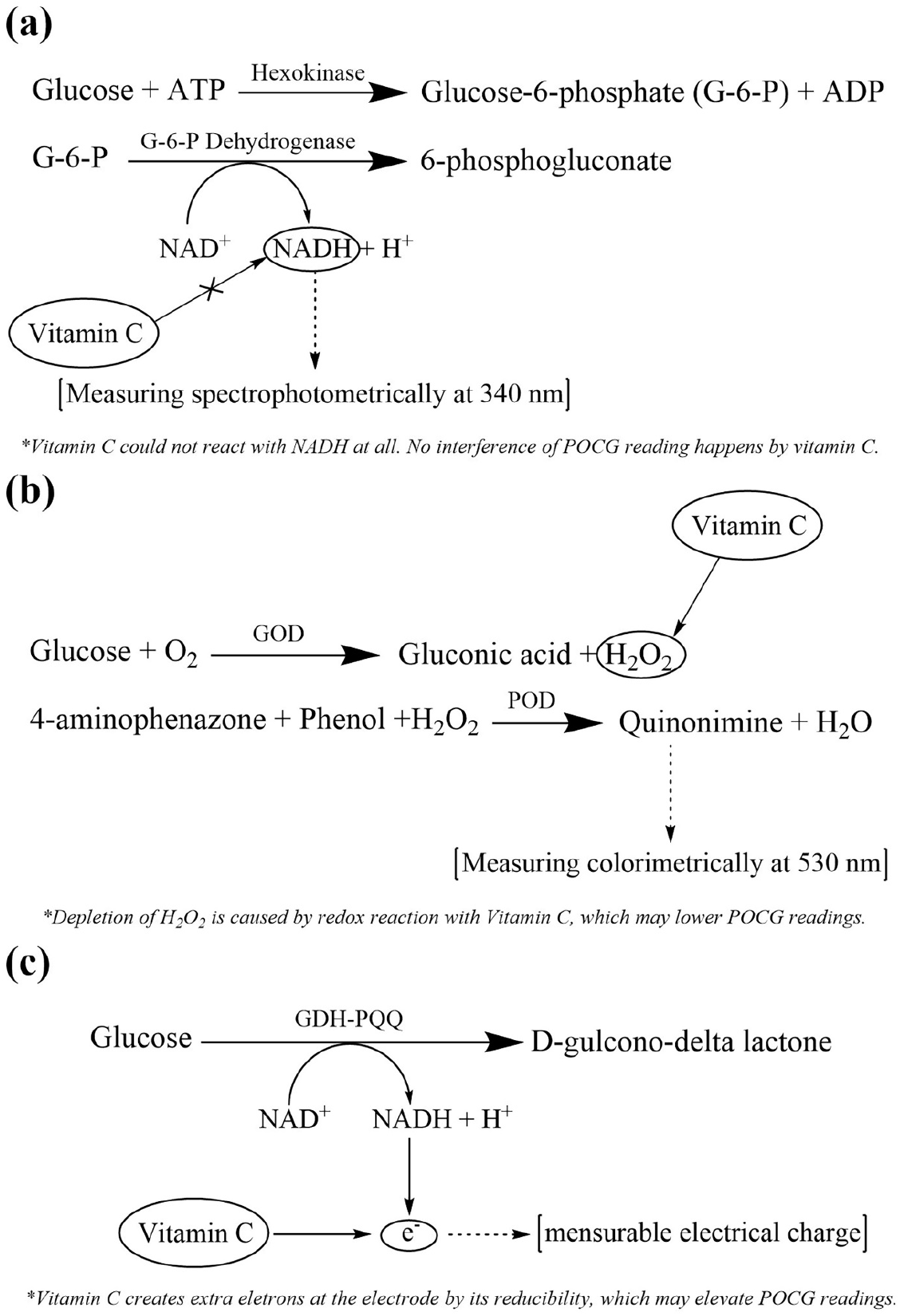
Three kinds of blood glucose monitoring methods: hexokinase spectrophotometric method (a); glucose oxidase-peroxidase (GOD-POD) colorimetric method (b); glucose dehydrogenase pyrroloquinoline quinone (GDH-PQQ) amperometric method (c).

GOD-POD colorimetric method is a convenient way of determining the level of glucose. GOD catalyzes the oxidation of glucose to gluconate, and hydrogen peroxide (H_2_O_2_) is liberated, which reacts with phenol and 4-aminophenazone (4-AP) in the presence of POD to form red quinonimine dye. The intensity of color was measured colorimetrically at 530 nm, which is directly proportional to the concentration of glucose in plasma [28, 29]. VC is a strong reductant and may set off redox reaction with H_2_O_2_, indicating that the depletion of H_2_O_2_ lowers the reading of POCG, which is consistent with the present research result (Figure 3b)[30].

Interestingly, our study exhibited a controversial result that POCG reading was lower than referential LG value. It was opposite to other researches while they showed incorrect POCG result could cause fictitious hyperglycemia by clinical measurement[25, 27, 31]. We found that other studies used glucose dehydrogenase pyrroloquinoline quinone (GDH-PQQ) amperometric method as POCG measurement method, which was different from GOD-POD colorimetric method by mechanism[25, 27, 31]. Nicotinamide adenine dinucleotide (NAD) is converted to NADH while Glucose is catalyzed to D-glucose-delta lactone by glucose dehydrogenase. NADH acts as an electron donor to create a mensurable electrical charge which could be quantified by an electrode (Figure 3c). Therefore, glucose level from blood sample is proportional to the sum of electron produced. As a strong reductant, VC is also an excellent electron donor which is oxidized at the electrode and release extra electrons to interfere the measurement result[25]. And that’s why fictitious hyperglycemia shows up when blood glucose measurement carries out by GDH-PQQ method in hdVC infusion period. This may indicate that higher or lower POCG erroneous reading may depend on various measuring methods when using hdVC as treatment. Since GDH-PPQ methods was widely utilized by plenty of blood glucose monitoring devices in Chinese hospitals, Fictitious lower POCG readings could become a potential medical risk induced by clinical application of hdVC diffusely.

Compliance with ISO15197:2013 criteria existed only 36.59% among all paired blood glucose values, which was far below the minimum accuracy criteria (95%). This also signified a great difference of compliance with ISO15197:2013 criteria among Smith (81.3%) and Howell’s researches (73.9%) respectively[27, 31]. Clinical influence of POCG value measuring interference by hdVC infusion was assessed by Parkes Consensus Error Grid Analysis in present study as well. Only 2.44% POCG value bias could cause significant medical risk when altering clinical action. In other words, inaccurate POCG reading by hdVC treatment could barely have a significant clinical effect in most situation.

According to some other researches, higher serum ascorbic acid concentration could be more likely to cause incorrect POCG reading[25, 26, 32, 33]. It depends on its daily dosage and renal excretion in vivo to a great extent. For further exploration, subgroup analysis was conducted to confirm if VC dosage or diverse level of patients’ renal function could be concernful affecting factors of erroneous POCG reading.

Although there is a consistency that more than 3g daily dosage of VC supplement is needed to maintain the normal serum normal serum vitamin C levels[34] and improve the clinical outcomes for severe ill patients[35, 36], the optimal dosage of VC as adjunctive treatment for sepsis is still unclear. Fowler *et al* finished a phase I trial for intravenous VC in 2014. Low dose (50mg/kg/24h) and high dose (200mg/kg/24h) were adopted for patients with severe sepsis every 6 hours[37]. Marik *et al* administered VC intravenously with 1.5g q6h with thiamine and hydrocortisone as the treatment of severe sepsis and septic shock[38]. Recently, the CITRIS-ALI Randomized Clinical Trial took 50mg/kg VC infusing every 6 hours for 4 days as treatment protocol[39]. With augmentation in serum VC concentration, POCG results may have a higher possibility to show an inaccurate reading. Hence, for the sake of verifying the extent of POCG reading interference by different VC dosage, we defined 50mg/kg/24h to 100mg/kg/24h as low dosage and 100mg/kg/24h to 200mg/kg/24h as high dosage respectively. Continuous infusion was adopted for maintaining more stable serum VC concentration to minimize bias as well. In the current study, higher VC dosage could induce POCG measurement bias easier than lower VC dosage. Significant medical risk could be more likely to generate in higher dosage group when altering clinical action in terms of erroneous POCG reading, compared to lower dosage group. It seems that higher dosage VC infusion could be more likely to induce unfaithful POCG measurement result. However, irrefutable conclusion could not be made due to the inadequate sample size of this research and lack of serum VC concentration monitoring, which should be taken into consideration in our further investigation.

Various levels of renal function for septic patients may also play an important role in excretion of VC in vivo due to its highly water-solubility and urinary elimination[8, 9]. Howell *et al* found that kidney function did not appeal to have a clinical impact on inaccurate POCG readings, but vasopressor administration interference was not excluded and patients with augmented renal clearance were not included in their research[31]. In terms of the Parkes Consensus Error Grid Analysis result in our study, there was an obviously larger difference between POCG and LG reading for patients who underwent CRRT treatment or suffered from impaired renal function. What’s more, significant clinical risk could not emerge when dealing with incorrect POCG value for patients with non-impaired renal function or augmented renal clearance, while 6.25% and 6.67% patients might have medical risk in impaired renal function and CRRT group respectively. Therefore, we could draw a conclusion that worse kidney function would cause POCG reading inaccuracy to greater extent. This fact is consistent with our hypothesis, which may be elaborated by VC accumulation in vivo due to decreasing renal elimination of VC. Similarly, affirmation could also not be made firmly because of uncertainty of VC serum concentration as well as comparatively small sample size.

The present study had some limitations. Firstly, it was a retrospective, single-center, observational case series with a small number of patients. Multiple-center prospective evaluation or randomized control trial (RCT) with a larger sample size should be put into practice. In addition to the GOD-POD method, other POCG measurement tools, such as glucose-dehydrogenase-pyrroloquinoline quinone (GOD-PQQ) amperometry method, were also available [25]. Taken together, further experiments along with other POCG measurement methods as replenishment are essential. As for the design of this study, it may be confused that researchers acquired blood glucose samples for paired measurement after hdVC infusion instead of during infusion period. On the one hand, since personalized hdVC treating protocols carried out as daily constant infusion with various dosages and days for different patients, researchers obtained blood samples after VC infusion for steady serum concentration uniformly. On the other hand, the 1.87-hour half-life time of intravenous VC in vivo owned minor effects when we got paired blood samples for measurement in 15 minutes after VC infusion[40]. Further researches should set up with a fixed treatment period and getting blood glucose samples during infusion time for contrast. Ascorbic acid concentration should be monitored as well. Another limitation for this research was lack of exclusion about other chemicals which could interfere accuracy of POCG measurement, such as glutathione, heparin, ibuprofen, etc[14]. Last but not the least, it is also important to ensure duration of hdVC treatment and monitor serum VC concentration during hdVC infusion period in future study.

## Conclusions

In summary, POCG measurement could be interfered by high-dose intravenous ascorbate acid infusion as compared to the reference LG method indeed. Although significant medical influence barely come up when altering clinical action by unreliable POCG reading, the LG method should be recommended to adopt for its accuracy. Moreover, elevating the dosage of VC would alter the POCG reading results to some extent. Renal function may also be another important affecting factor as well. Further investigations such as RCTs or prospective trials should be put into practice to verify the results of the current study.

## Data Availability

The datasets used and/or analyzed during the current study are available from the corresponding author on reasonable request.

## Transparency

### Declaration of Funding

This study was supported by Shanghai Shen Kang Hospital Development Center Clinical Science and technology innovation project (No. SHDC12017116) and Important and weak discipline construction plan for health and family planning system of Shanghai (No. 2016ZB0206).

### Declaration of financial/other relationships

All authors declared no financial or other relationships in this study should be declared in this paper.

### Author contributions

GZ, JH and XB had full access to all data in the study and take responsibility for the integrity and the accuracy of the data analysis; they contributed equally to the manuscript. GZ, JH and XB conceived and designed this study, participated in study design and coordination, and helped to draft the manuscript. GZ, JH, XQ, BZ, HS, XB, BC, EC and EM collected the information, and contributed to the acquisition, analysis and interpretation of the data. GZ, JH and XB wrote and revised the manuscript. All authors read and approved the final manuscript.

## Acknowledgements

The authors thank the staff of the EICU Department of Ruijin Hospital affiliated to Medical School of Shanghai Jiao Tong University for their facilities and collaboration.

## Notes

### Competing Interest Statement

The authors have declared no competing interest.

### Author Declarations

This study was approved by Ruijin Hospital Institutional Review Board and has been performed in accordance with the ethical standards laid down in Declaration of Helsinki 1964 and its later amendments or comparable ethical standards.

